# The germline DNA repair gene landscape in *BRCA1* and *BRCA2* implicated hereditary breast cancer families

**DOI:** 10.1101/2025.06.24.25329575

**Authors:** Wejdan M. Alenezi, Neil Recio, Caitlin T. Fierheller, Corinne Serruya, Timothée Revil, Anne-Marie Mes-Masson, Diane Provencher, Celia M.T. Greenwood, Jiannis Ragoussis, Patricia N. Tonin

## Abstract

As gene-panels expand to include candidate breast cancer predisposing genes (CPGs) involved in diverse DNA repair pathways, we investigated an index breast cancer (BC) case from 56 *BRCA1* or *BRCA2* implicated high-risk hereditary BC families for rare germline variants predicted to be damaging and clinically relevant in 276 DNA repair genes (DRGs). Using whole exome sequencing analyses, we identified a total of 287 variants in 55% of DRGs, of which 24 loss-of-function and 36 other predicted damaging variants were identified in 72% and 76% of *BRCA1* and *BRCA2* implicated cases, respectively. We also identified 60 variants of clinical interest in various known CPGs, including those involved in risk to other cancers, in 72% of *BRCA1* and 60% of *BRCA2* cases. The number of variants predicted to be deleterious in diverse DRGs raises questions about the interpretability of findings as panel testing expands beyond clinically established breast CPGs.

## INTRODUCTION

Heterozygous carriers of germline pathogenic variants (PVs) in *BRCA1* or *BRCA2* have a high life-time risk for female breast cancer (BC) estimated at 55-72%, depending on the gene involved^1^. PVs in these major BC predisposing genes (BCPGs) have been implicated in a significant proportion of Hereditary Breast and/or Ovarian Cancer (HBC/HBOC) syndrome families which exhibit an autosomal dominant mode of inheritance^1^. Germline genetic testing identifies carriers of PVs in *BRCA1* or *BRCA2* who may benefit from cancer screening, prevention and therapeutic management strategies^1,2^. Other BCPGs implicated in HBC syndrome families include *ATM, BARD1, CHEK2, PALB2, RAD51C* and *RAD51D*^3,4^. Though *CDH1, NF1, PTEN, STK11* and *TP53* are involved in high-risk BC, these CPGs have been associated with cancer syndromes distinct from HBC and HBOC^1,3,4^. Though all of these BCPGs have been included in gene-panel tests for BC risk assessment and management, *BRCA1* and *BRCA2* remain the major risk genes accounting for about one-third of all HBC families, with other BCPGs collectively accounting for less than 5% of such families^3,5^.

With the increased use of gene-panel testing and sequencing-based screening, there have been reports of rare carriers of PVs in at least two BCPGs^6–8^. Although the implications are unknown for BC risk, phenotype or disease progression, these observations are intriguing given that the established BCPGs play critical roles in safeguarding the genome through DNA damage response repair^9^. Prompted by our earlier report of the co-occurrence of PVs in both *PALB2* and *BRCA1* in a familial BC case of French Canadian (FC) ancestry^10^, we used targeted-variant analysis in index carriers of PVs in *BRCA1* or *BRCA2* that were selected from high-risk BC families^11,12^. The FC of Quebec Canada exhibit a significant frequency of specific PVs in *BRCA1, BRCA2, PALB2* and *RAD51D* due to genetic drift, a genetic architecture that has facilitated the interpretation of rare variants identified in candidate hereditary cancer risk genes applicable to other populations^13–16^. Here, we report the analyses of whole exome sequencing (WES) to investigate the genetic landscape of 276 DNA repair genes (DRGs)^17^, which include the established BCPGs and CPGs implicated in other hereditary cancers.

## RESULTS

### Rare predicted damaging variants

Rare variants in 276 DRGs^17^ were identified in WES data from 18 *BRCA1* and 38 *BRCA2* implicated BC (*BRCA1/BRCA2*) cases from HBC families using a previously established variant selection and annotation pipeline^18^ (**Methods; Supplementary Tables 1, 2**). A total of 571 variants were identified with minor allele frequencies (MAFs) ≤0.01 or were not reported in The Genome Aggregation Database v.2.1.1. (gnomAD), where 485 had read depths ≥10 and variant allele frequencies (VAFs) ≥0.2. All PVs in the 56 *BRCA1*/*BRCA2* cases were re-identified in our WES data and excluded from further analyses. From the remaining total of 377 variants that met our filtering criteria, 287 were unique predicted damaging variants by at least 1 out of 12 best performing in silico tools^19^ (**Supplementary Table 3**) and analyzed further.

The 287 unique variants including newly detected variants in *BRCA1* and *BRCA2* were identified in 55% (153/276) of DRGs involved in diverse pathways^17^: Base Excision Repair (BER), Fanconi Anemia (FA), Homologous Recombination (HR), Non-Homologous End Joining (NHEJ) and Mismatch Repair (MMR) (**Supplementary Table 3**). All 56 *BRCA1*/*BRCA2* cases carried 2 to 13 (median 6) of these unique variants. All but 6 (2%; 6/287) variants were identified in a heterozygous state. About 30% (87/287) of variants had MAFs of ≤10^−4^ which are as rare as MAF of PVs in *BRCA1* and *BRCA2* reported in the general population according to gnomAD, and about 10% (30/287) have not been reported in this genetic database. These 117 rare variants were identified in 34% (94/276) of DRGs, and were found in 94% (17/18) of *BRCA1* and 92% (35/38) of *BRCA2* cases. About 8% (24/287) of variants were predicted to incur loss-of-function (LoF): 11 frameshift, 9 stop-gain, 3 canonical splice-site (delta score ≥0.5 by SpliceAI^20^) and 1 start-lost variant. About 39% (7/18) of *BRCA1* and 50% (19/38) of *BRCA2* cases carried 1 to 2 of these 24 LoF variants. The remaining 92% (263/287) of variants were predicted to have a moderate effect: 226 missense, 25 non-canonical splice-site, 7 in-frame deletions and 5 synonymous variants. Among these 263 variants, 32 missense variants were redicted to be damaging by REVEL (score ≥0.7)^21^, and 3 non-canonical splice site and 1 synonymous variant were predicted to effect splicing by SpliceAI. One to 4 of these 36 predicted damaging variants were identified in 50% (9/18) of *BRCA1* and 45% (17/38) of *BRCA2* cases. Therefore, of the total of LoF and predicted damaging variants, 21% (60/287) were identified in 72% (13/18) of *BRCA1* and 76% (29/38) of *BRCA2* cases, where each carried 1 to 5 (median 2) of any these variants.

About 7% (21/287) of unique variants were identified in 9/19 established BCPGs^1^. These included variants in *ATM, BARD1, BRCA1, BRCA2, CHEK2, PALB2, RAD51C, RAD51D* and *TP53*, 9 of which had MAFs ≤10^−4^. They were identified in 33% (6/18) of *BRCA1* and 39% (15/38) of *BRCA2* cases. There were 3 LoF (*ATM* c.3802del; p.Val1268Ter, *BRCA2* c.9976A>T; p.Lys3326Ter and *CHEK2* c.247del; p.Gln83LysfsTer54), 1 non-canonical splice-site and 17 missense variants. Three of these variants were predicted to be damaging: *BRCA2* c.4585G>A; p.Gly1529Arg, *BARD1* c.1339C>G; p.Leu447Val and *CHEK2* c.667C>T; p.Arg223Cys.

### Clinical classification of rare variants

To infer the clinical consequences of carrying damaging variants in DRGs, the 287 unique variants were classified according to The American College of Medical Genetics and Genomics (ACMG) guidelines^22^ (**Methods**; **Supplementary Table 3**). A total of 60 variants of clinical interest 5 PVs, 10 likely pathogenic (LPVs) and 45 variants of uncertain significance (VUSs) were identified in 18% (51/276) of DRGs (**Figure 1**). They were identified in 72% (13/18) of *BRCA1* and 60% (23/38) of *BRCA2* cases, where each case carried 1 to 4 (median of 2) of such variants. Many of these variants (93%; 56/60) had MAFs ≤10^−4^ or have not been reported in gnomAD.

**Figure 1.**
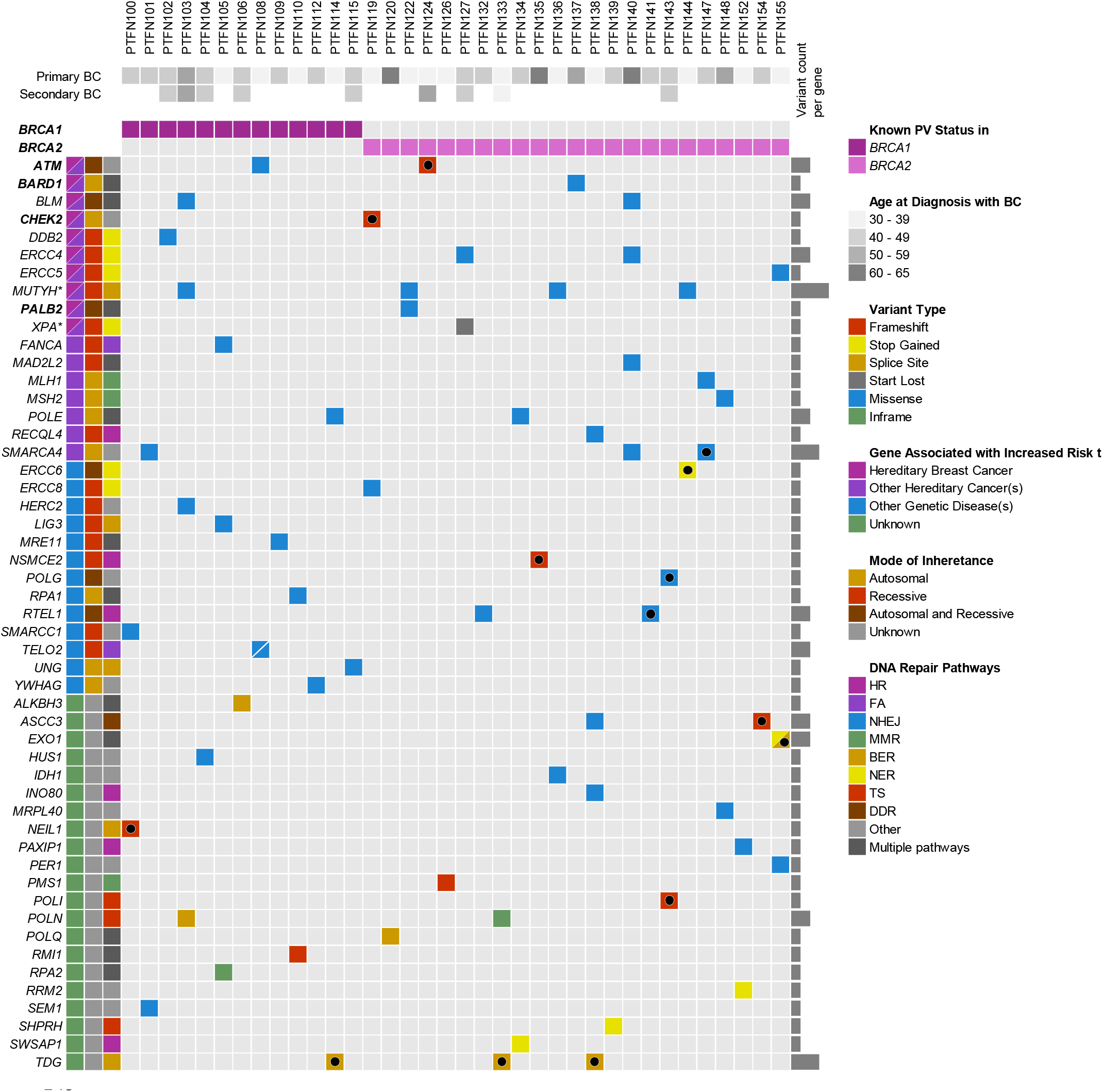
Oncoprint of 60 predicted damaging variants of clinical interest identified in *BRCA1* and *BRCA2* implicated index BC cases. Genes in bold are established breast cancer (BC) predisposing genes according to the National Comprehensive Cancer Network Clinical Practice in Oncology Guidelines (Version 3.2025)—Genetic/Familial High-Risk Assessment: Breast, Ovarian and Pancreatic^1^. Variants classified by The American College of Medical Genetics and Genomics (ACMG) guidelines^22^ (**Methods**; **Supplementary Table 3**) indicated with colour coded square (based on variant type) where black dot indicates pathogenic (PV) or likely pathogenic variant (LPV), and all other colour coded squares as variants of uncertain clinical significance (VUS). Variants in genes indicated with an asterisk (*) were identified in the heterozygous state, while ACMG guidelines classify variants in these genes as pathogenic when present in the homozygous state. To protect anonymity of participants approximate ages of diagnoses of cancers a provided to the nearest 10th-year age range at diagnoses.

About 8% (5/60) of variants of clinical interest were identified in 5 different BCPGs. These include *ATM* c.3802del; p.Val1268Ter and *CHEK2* c.247del; p.Gln83LysfsTer54 that conferred LoF and were classified as PV/LPVs and 3 missense variants in *ATM, BARD1* or *PALB2*, which were classified as VUSs. These variants were identified in 6% (1/18) of *BRCA1* and 11% (4/38) of *BRCA2* cases. There were 16 other variants in 9 BCPGs that were classified as benign or likely benign involving *BRCA1, BRCA2, RAD51C, RAD51D, CHEK2, PALB2, RAD51C, RAD51D* and *TP53*. Among these variants, two were LoF: *BRCA2* c.9976A>T; p.Lys3326Ter and *CHEK2* c.320-5T>C, and two were missense *BRCA2* c.4585G>A; p.Gly1529Arg and *CHEK2* c.667C>T; p.Arg223Cys, which were predicted to be damaging by REVEL.

About 11% (7/60) of variants of clinical interest were identified in 5 established CPGs involved in other types of hereditary cancer. Three VUSs were identified in a heterozygous state in 4 BC cases: *MLH1* c.415C>G; p.Pro139Ala or *MSH2* c.2118C>G; p.Asp706Glu and *POLE* c.320C>T; p.Ala107Val. Two PV and LPV: *MUTYH* c.1178G>A; p. Gly393Asp, c.691G>A; p.Val231Met and c.1178G>A; p.Gly393Asp. Three BC cases were identified carrying a LPV c.2858A>G; p.Lys953Arg or a VUS c.919C>T; p.Pro307Ser in *SMARCA4*. None of carriers of these variants reported a personal or family history of cancer predictive of the involvement of any of these CPGs (**Supplementary Table 2**).

The remaining 80% (48/60) of variants of clinical interest were identified in 42 different DRGs that have not been associated with cancer risk. They were identified in 72% (13/18) of *BRCA1* and 50% (19/38) of *BRCA2* cases. They include 9 PVs/LPVs in *ASCC3, ERCC6, EXO1, NEIL1, NSMCE2, POLG, POLI, RTEL1* and *TDG* that were identified in 11% (2/18) of *BRCA1* and 21% (8/38) of *BRCA2* cases, where there were 3 carriers of *TDG* c.1090+1G>T. There was one heterozygous carrier of *XPA* c.3G>T; p. Met1, which was classified as PV in the homozygous state. Carriers of VUSs in *ASCC3, EXO1* and *RTEL1* were also identified as were 2 carriers each of VUSs in *BLM, ERCC4, POLN, RTEL1* and *TELO2*.

## DISCUSSION

Our analyses of WES data from 56 *BRCA1*/*BRCA2* BC cases from HBC families identified 287 unique variants predicted to be damaging in 153 of 276 (55%) diverse DRGs. Damaging or clinically relevant variants were identified in the other established BCPGs^1^: *ATM, BARD1, CHEK2, PALB2, RAD51C, RAD51D* and *TP53*, but not in *PTEN*. Variants of clinical interest were also identified in *POLE, MLH1* and *MSH2*, CPGs involved in hereditary colorectal, endometrial and ovarian cancers^23^, and in *SMARCA4*, a CPG involved in hereditary small cell carcinoma of the ovary, hypercalcemic type and rhabdoid tumour^24^, though both personal and family history of cancer from index carriers would not necessarily predict involvement of these CPGs. Interestingly, heterozygous carriers of PV/LPVs in *MUTYH* were identified, a CPG involved in the autosomal recessive form of hereditary colorectal cancer^23^. Though monoallelic *MUTYH* variants have been reported in BC cases^23,25^ where one study estimated a 1.4-fold increased risk for BC among carriers^26^ the findings are not equivocal^23,27,28^. Our findings suggest that BC cases from high-risk BC families harbouring PVs in *BRCA1* or *BRCA2* may also carry clinically relevant variants in other established CPGs.

With increasing use of gene panel testing are reports of rare cancer cases harbouring PVs in more than one clinically relevant CPG. Multi-locus inherited neoplasia syndrome (MINAS) refers to the co-occurrence of PVs in different CPGs^6^. Recently, the mean age at BC diagnosis in co-occurring *BRCA1* and *BRCA2* PV carriers was found similar or moderately younger than carriers with a *BRCA1* PV alone though findings were limited due to sample size^29^. As with our findings, co-occurring PVs in *BRCA1*/*BRCA2* and other established BCPG^6,29^ such as *ATM* or *CHEK2* were observed. A recent analysis of the BRIDGES dataset for variants of interest in 11 BCPGs among 55,815 BC cases, reported that the co-occurrence of PVs in *BRCA1, BRCA2* and *PALB2* was less frequent than expected compared to the other BCPGs analyzed^30^. The authors suggested that the co-occurrence of VUSs in *BRCA1, BRCA2* or *PALB2* with PVs in other BCPGs may provide supportive evidence against pathogenicity of those VUSs^30^.

Our study also identified variants of clinical interest in DRGs that are not established CPGs, such as PV/LPVs in *ASCC3, ERCC6, EXO1, NEIL1, NSMCE2, POLG, POLI, RTEL1, TDG* and *XPA* based on ACMG guidelines. Genetic studies have implicated PVs in *ERCC6* and *EXO1* in hereditary BC cases not accounted for by *BRCA1*/*BRCA2*^31–34^. Interestingly, specific missense variants in *POLG* and *RTEL1* have been reported two or more times more frequently in BC cases as compared to controls^35,36^.

Our findings suggest that the frequency of BC carriers with clinically relevant co-occurring alleles in *BRCA1*/*BRCA2* and other BCPGs may be higher than expected. One large population-based study reported that 5.03% of BC cases versus 1.63% of controls carry PV in any of 12 established BCPGs though this carrier frequency may be higher in ancestrally defined populations^3^. Though reports of recurrent, founder PVs in *BRCA1, BRCA2, CHEK2, PALB2* and *TP53* in FC BC cases not selected for family history of BC revealed carrier frequencies ranging from 0.3% to 6.4% versus <1% of controls depending on the BCPG investigated^15,37,38^, we are only aware of our report of a single familial BC case harbouring PVs in both *BRCA1* and *PALB2* from a FC family^10^. With respect to other DRGs investigated in our study, it is interesting that our group also reported different PVs in *EXO1* and *NEIL1* in WES data from familial ovarian cancer cases of FC ancestry not accounted for by *BRCA1* and *BRCA2*^18^.

An important consideration in the interpretation of the variant burden in our study cohort is the potential influence of Berkson’s paradox, so-called collider bias^39^. As our BC cases were selected based on having a strong family history of BC diagnosed in close relatives in the same familial branch, there exists an inherent selection bias favouring individuals who are more likely to carry additional rare germline variants in other genes. This non-random sampling may artificially inflate the observed co-occurrence of *BRCA1* and *BRCA2* PVs with other clinically relevant variants in DRGs that were identified in our study, potentially leading to a spurious inverse or exaggerated association between these variants and other genetic or family history features. Recognizing this phenomenon is critical for contextualizing variant enrichment and highlights the need for cautious interpretation of genetic interactions in highly selected hereditary cancer study groups.

Questions remain concerning the biological significance of harbouring more than one PV in established BCPGs. Studies of heterozygous carriers with co-occurring PVs in *BRCA1* and *BRCA2* suggested that BC tumours most likely develop from the loss of the functioning allele but not both implying that the biological effects at the somatic level are additive rather than synergistic^6^. BC is a heterogeneous disease with a high degree of somatic molecular genetic diversity both between and within tumors which together with the heritable germline landscape likely affects disease progression and sensitivity to chemo- and radiotherapy beyond the impact of carrying a CPG risk allele^40^. With respect to our findings, several reports have suggested that variants in or near *ERCC6, EXO1, NSMCE2*, and *POLG* may be clinically relevant to BC. *ERCC6* SNPs may modify occupational radiation exposure and BC risk^41^. Increased *EXO1* expression has been associated with breast carcinogenesis and poor prognosis^42^. A newly developed Capture Hi-C assay identified protein-coding genes including *NSMCE2* as potential targets of genome-wide association analyses identified loci associated with BC risk^43^. Interestingly, *NSMCE2*, a super-enhancer regulated gene, when expressed at high levels was linked to poor prognosis and therapy resistance in BC^44^. Similarly, SNPs in *POLG* have been associated with tumour phenotype and disease outcome in BC patients^45^.

Notably, the participants in our study group could have benefited from poly(ADP-ribose) polymerase (PARP) inhibitor therapy since they carry PVs in *BRCA1*/*BRCA2* that affect the HR pathway. The clinical implications for participants who also carried variants in DRGs involved in other pathways is unknown. An investigation of an ovarian cancer cell line model system suggested that responses to PARP inhibitors may be due to deficiencies in DRGs in defined pathways such as HR and either NER or MMR^46^. Further research is needed on the vulnerabilities of BC tumours arising in carriers of co-occurring PVs affecting functionally diverse DRGs.

## CONCLUSION

Our analysis of 56 index BC cases with *BRCA1*/*BRCA2* PVs from high-risk HBC families, with shared ancestry, identified carriers of co-occurring, predicted damaging and clinically relevant alleles in DRGs. The frequency of these carriers may be higher than expected and warrants further investigation of other populations at high risk for hereditary BC. The number of predicted damaging and clinically relevant variants identified in different DRGs raises questions concerning the interpretability of findings as panel testing expands beyond established BCPGs.

## METHODS

### Study subjects

The 56 index BC cases for WES analyses were selected from 56 high-risk BC families that have been extensively characterized by our group and previously reported to carry PVs in *BRCA1* (n=18) or *BRCA2* (n=38)^11,12^ (**Supplementary Table 2**). Briefly, index cases were diagnosed with BC <66 years of age and had a family history of at least two confirmed cases of invasive BC diagnosed <66 years of age in first-, second-, or third-degree relatives in the same familial branch by review of pathology reports or death certificates; and reported FC of Quebec ancestry for all four grandparents as described previously^11,12^. The ages of diagnosis of BC and personal history of other cancer types in 10-year intervals to protect anonymity of participants are described in **Supplementary Table 2**. Peripheral blood lymphocyte (PBL) DNA samples, ages of diagnoses of personal and family history of cancer were available from index cases from an established biobank resource: the Banque de tissus et données of the Réseau de recherche sur le cancer of the Fond de recherche du Québec – Santé (RRCancer biobank) (rrcancer.ca/) where participants had been recruited in adult hereditary cancer clinics in accordance with ethical guidelines of their Research Ethics Boards. This project was conducted in accordance with the guidelines of The McGill University Health Centre Research Ethics Board.

### WES analysis of DNA repair genes

WES was performed using PBL DNA from the 56 index BC cases at the McGill Genome Centre and rare germline variants were identified using a previously described variant selection and annotation pipeline^18^. In brief, for primary and secondary analysis, NimbleGen SeqCap^®^ EZ Exome v3.0 library kit (Roche, US), followed by paired-end sequencing on different Illumina HiSeq platforms was performed. Reads were aligned to the human reference genome assembly GRCh37/hg19 using Burrows-Wheeler aligner v0.7.17, followed by PCR deduplication using Picard v2.9.0. Realignment around small insertions and deletions was performed, and germline variants were called using HaplotypeCaller using Genome Analysis Toolkit (GATK) v3.5. Variants were then filtered for base sequencing quality score ≥30 and annotated using Ensembl Variant Effect Predictor (VEP) and GEMINI v0.19.1.

For tertiary analysis, the list of annotated variants were filtered for those: (1) occurring in protein coding and splice-site regions of 276 DRGs^17^ including *BRCA1* and *BRCA2* (**Supplementary Table 1**); (2) having a MAF ≤0.01 in the general population based or not reported in gnomAD (gnomad.broadinstitute.org/); and (3) having a read depth ≥10 and variant allele frequency (VAF) ≥0.2. The resulting list of 470 unique variants, excluding the 15 known PVs in the 56 *BRCA1* or *BRCA2* cases, were annotated using 8 selected in silico tools according to their predictions for being damaging (Combined Annotation Dependent Depletion v1.4 (CADD [Phred score ≥20]), Eigen (score ≥0.0), Meta-analytic Logistic Regression v4.2 (MetaLR [score ≥0.5]), Meta-Analytic Support Vector Machine v4.2 (MetaSVM [score ≥0.0]), MetaRNN 4.2 (score ≥0.5), Rare Exome Variant Ensemble Learner v4.2 (REVEL [score ≥0.7)), Variant Effect Scoring Test v4.2 (VEST [score ≥0.5]) and Protein Variation Effect Analyzer v4.0 (PROVEAN v4.0 [score ≤−2.5]) or four selected tools for affecting splicing (Maximum Entropy Estimates of Splice Junction v2.0 (MaxEntScan [score≥0.5]), Database Splicing Consensus Single Nucleotide Variant (dbscSNV): AdaBoost v4.0 (ADA [score ≥0.5]) and Random Forest v4.0 (RF [score≥0.5]) and SpliceAI (score ≥0.5) using in silico tools that were selected for their best performance as previously described^19^. The final list of 287 unique variants, which were predicted to be damaging by at least one of these in silico tools, were classified as PV, LPV, VUS, benign or likely benign based on the ACMG^22^ guidelines as previously described^18^. This list of 287 variants was verified manually using the Integrative Genomics Viewer (IGV) v2.8. (igv.org).

## DATA AVAILABILITY

Data supporting the findings presented in this study are available within the article and its supplementary material. Selected data will also be available in the Leiden Open Variation Database (LOVD^3^): Disease #04296 - Neoplasia Multiple Inherited Alleles (MINAS) database (databases.lovd.nl/shared/diseases/04296).

## ACKNOWLEDGEMENTS

This research study was made possible with DNA samples and data from cancer families through the collaboration with the Réseau de recherche sur le cancer (RRCancer) which is affiliated to the Canadian Tissue Repository Network (CTRNet); Suzanna Arcand for preparing samples for whole exome sequencing (WES); and Frédéric Ancot for targeted analyses of selected genes in original data set.

## Funding

W.M.A. was supported by a Scholarship Award and The Ministry of Education, Saudi Arabia. N.R was supported by the Fonds recherche Québec Santé (FRQS) scholarship award. C.T.F. was supported by The Research Institute of the McGill University Health Centre Studentship (RI-MUHC) Award and Ovarian Cancer Canada Trainee Travel (Research) Award, and James O. and Maria Meadows Award. Research was supported by operating grants from Cancer Research Society operating grant (Number 21123 to P.N.T.) and Canadian Institute for Health Research (CIHR) (PJT-156124 to P.N.T., J.R.); FRQS and Breast Cancer Foundation infrastructure grants to RRCancer of the FRQS (to P.N.T). The RI MUHC and the Centre hospitalier de l’Université de Montréal receive support from the FRQS.

## AUTHOR CONTRIBUTIONS

W.M.A. performed tertiary bioinformatics analysis of WES data. N.R. and C.F. assisted in the bioinformatics analysis of WES data. T.R. performed primary and secondary bioinformatics analysis of WES data. C.S. assisted in verifying the clinical data. A.-M. M.-M, and D.P. assisted in identifying cases from biobanks. C.M.T.G. oversaw the statistical analyses. J.R. and P.N.T oversaw the WES data collection and analyses. W.M.A drafted the initial versions of the manuscript. W.M.A. and P.N.T. designed and supervised all aspects of the study and edited the manuscript. All authors reviewed and edited the final version of the manuscript.

## COMPETING INTERESTS

Authors declare that there are no conflicts of interest.

## ETHICS APPROVAL

This study was approved by the Research Ethics Board of the Research Centre of the McGill University Health Centre, Montreal, Canada (MP-37-2019-4783).

## Notes

### Competing Interest Statement

The authors have declared no competing interest.

### Funding Statement

WMA was supported by a Scholarship Award and The Ministry of Education Saudi Arabia NR was supported by the Fonds recherche Quebec Sante FRQS scholarship award. CTF was supported by The Research Institute of the McGill University Health Centre Studentship RIMUHC Award and Ovarian Cancer Canada Trainee Travel Research Award and James O and Maria Meadows Award Research was supported by operating grants from Cancer Research Society operating grant to PNT and Canadian Institute for Health Research CIHR to PNT JR FRQS and Breast Cancer Foundation infrastructure grants to RRCancer of the FRQS to PNT The RI MUHC and the Centre hospitalier de le Universite de Montreal receive support from the FRQS

### Author Declarations

This study was approved by the Research Ethics Board of the Research Centre of the McGill University Health Centre Montreal Canada

## REFERENCES

1. Daly, M. B. et al. NCCN Guidelines® Insights: Genetic/Familial High-Risk Assessment: Breast, Ovarian, and Pancreatic, Version 2.2024. J Natl Compr Canc Netw 21, 1001– 1010 (2023).

2. Bedrosian, I. et al. Germline Testing in Patients With Breast Cancer: ASCO-Society of Surgical Oncology Guideline. J Clin Oncol 42, 584–604 (2024).

3. Hu, C. et al. A Population-Based Study of Genes Previously Implicated in Breast Cancer. New England Journal of Medicine 384, 440–451 (2021).

4. Breast Cancer Risk Genes — Association Analysis in More than 113,000 Women. New England Journal of Medicine 384, 428–439 (2021).

5. Slavin, T. P. et al. The contribution of pathogenic variants in breast cancer susceptibility genes to familial breast cancer risk. NPJ Breast Cancer 3, (2017).

6. McGuigan, A. et al. Multilocus Inherited Neoplasia Allele Syndrome (MINAS): an update. Eur J Hum Genet 30, 265–270 (2022).

7. Laitman, Y. et al. Phenotypes of carriers of two mutated alleles in major cancer susceptibility genes. Breast Cancer Res Treat 208, (2024).

8. Infante, M. et al. Increased Co-Occurrence of Pathogenic Variants in Hereditary Breast and Ovarian Cancer and Lynch Syndromes: A Consequence of Multigene Panel Genetic Testing? Int J Mol Sci 23, 11499 (2022).

9. Nielsen, F. C., Van Overeem Hansen, T. & Sørensen, C. S. Hereditary breast and ovarian cancer: new genes in confined pathways. Nat Rev Cancer 16, 599–612 (2016).

10. Ancot, F., Arcand, S. L., Mes-Masson, A. M., Provencher, D. M. & Tonin, P. N. Double PALB2 and BRCA1/BRCA2 mutation carriers are rare in breast cancer and breast-ovarian cancer syndrome families from the French Canadian founder population. Oncol Lett 9, 2787–2790 (2015).

11. Oros, K. K. et al. Application of BRCA1 and BRCA2 mutation carrier prediction models in breast and/or ovarian cancer families of French Canadian descent. Clin Genet 70, 320– 329 (2006).

12. Cavallone, L. et al. Comprehensive BRCA1 and BRCA2 mutation analyses and review of French Canadian families with at least three cases of breast cancer. Fam Cancer 9, 507– 517 (2010).

13. Scriver, C. R. Human genetics: lessons from Quebec populations. Annu Rev Genomics Hum Genet 2, 69–101 (2001).

14. Laberge, A. M. et al. Population history and its impact on medical genetics in Quebec. Clin Genet 68, 287–301 (2005).

15. Fierheller, C. T., Alenezi, W. M. & Tonin, P. N. The Genetic Analyses of French Canadians of Quebec Facilitate the Characterization of New Cancer Predisposing Genes Implicated in Hereditary Breast and/or Ovarian Cancer Syndrome Families. Cancers (Basel) 13, (2021).

16. Anderson-Trocmé, L. et al. On the genes, genealogies, and geographies of Quebec. Science 380, 849–855 (2023).

17. Knijnenburg, T. A. et al. Genomic and Molecular Landscape of DNA Damage Repair Deficiency across The Cancer Genome Atlas. Cell Rep 23, 239-254.e6 (2018).

18. Alenezi, W. M. et al. Genetic analyses of DNA repair pathway associated genes implicate new candidate cancer predisposing genes in ancestrally defined ovarian cancer cases. Front Oncol 13, (2023).

19. Cubuk, C. et al. Clinical likelihood ratios and balanced accuracy for 44 in silico tools against multiple large-scale functional assays of cancer susceptibility genes. Genet Med 23, 2096–2104 (2021).

20. Jaganathan, K. et al. Predicting Splicing from Primary Sequence with Deep Learning. Cell 176, 535-548.e24 (2019).

21. Ioannidis, N. M. et al. REVEL: An Ensemble Method for Predicting the Pathogenicity of Rare Missense Variants. Am J Hum Genet 99, 877–885 (2016).

22. Richards, S. et al. Standards and guidelines for the interpretation of sequence variants: a joint consensus recommendation of the American College of Medical Genetics and Genomics and the Association for Molecular Pathology. Genet Med 17, 405–424 (2015).

23. Gupta, S. et al. NCCN Guidelines Version 3.2024 Genetic/Familial High-Risk Assessment: Colorectal, Endometrial, and Gastric Continue. (2024).

24. Witkowski, L. et al. Germline and somatic SMARCA4 mutations characterize small cell carcinoma of the ovary, hypercalcemic type. Nature Genetics 2014 46:5 46, 438–443 (2014).

25. Van Der Kolk, D. M. et al. Penetrance of breast cancer, ovarian cancer and contralateral breast cancer in BRCA1 and BRCA2 families: high cancer incidence at older age. Breast Cancer Res Treat 124, 643–651 (2010).

26. Win, A. K. et al. Risk of extracolonic cancers for people with biallelic and monoallelic mutations in MUTYH. Int J Cancer 139, 1557–1563 (2016).

27. Out, A. A. et al. MUTYH gene variants and breast cancer in a Dutch case–control study. Breast Cancer Res Treat 134, 219–227 (2012).

28. Fulk, K. et al. Monoallelic MUTYH carrier status is not associated with increased breast cancer risk in a multigene panel cohort. Fam Cancer 18, 197–201 (2019).

29. Laitman, Y. et al. Phenotypes of carriers of two mutated alleles in major cancer susceptibility genes. Breast Cancer Res Treat 208, 589–595 (2024).

30. Davidson, A. L. et al. Co-observation of germline pathogenic variants in breast cancer predisposition genes: Results from analysis of the BRIDGES sequencing dataset. Am J Hum Genet 111, (2024).

31. Jalkh, N. et al. Next-generation sequencing in familial breast cancer patients from Lebanon. BMC Med Genomics 10, 1–12 (2017).

32. Merdad, A. et al. Characterization of familial breast cancer in Saudi Arabia. BMC Genomics 16 Suppl 1, (2015).

33. Kurkilahti, V. et al. Rare Germline Variants in DNA Repair Genes Detected in BRCA-Negative Finnish Patients with Early-Onset Breast Cancer. Cancers (Basel) 16, 2955 (2024).

34. Shahi, R. B. et al. Identification of candidate cancer predisposing variants by performing whole-exome sequencing on index patients from BRCA1 and BRCA2-negative breast cancer families. BMC Cancer 19, (2019).

35. Tervasmäki, A. et al. Rare missense mutations in RECQL and POLG associate with inherited predisposition to breast cancer. Int J Cancer 142, 2286–2292 (2018).

36. Girard, E. et al. Familial breast cancer and DNA repair genes: Insights into known and novel susceptibility genes from the GENESIS study, and implications for multigene panel testing. Int J Cancer 144, 1962 (2018).

37. Ghadirian, P. et al. The contribution of founder mutations to early-onset breast cancer in French-Canadian women. Clin Genet 76, 421–426 (2009).

38. Behl, S. et al. Founder BRCA1/BRCA2/PALB2 pathogenic variants in French-Canadian breast cancer cases and controls. Sci Rep 10, (2020).

39. Westreich, D. Berksons bias, selection bias, and missing data. Epidemiology 23, 159– 164 (2012).

40. Lüönd, F., Tiede, S. & Christofori, G. Breast cancer as an example of tumour heterogeneity and tumour cell plasticity during malignant progression. British Journal of Cancer 2021 125:2 125, 164–175 (2021).

41. Rajaraman, P. et al. Nucleotide excision repair polymorphisms may modify ionizing radiation-related breast cancer risk in US radiologic technologists. Int J Cancer 123, 2713–2716 (2008).

42. Liu, J. & Zhang, J. Elevated EXO1 expression is associated with breast carcinogenesis and poor prognosis. Ann Transl Med 9, 135–135 (2021).

43. Dryden, N. H. et al. Unbiased analysis of potential targets of breast cancer susceptibility loci by Capture Hi-C. Genome Res 24, 1854–1868 (2014).

44. Di Benedetto, C. et al. NSMCE2, a novel super-enhancer-regulated gene, is linked to poor prognosis and therapy resistance in breast cancer. BMC Cancer 22, (2022).

45. Golubickaite, I. et al. The impact of mitochondria-related POLG and TFAM variants on breast cancer pathomorphological characteristics and patient outcomes. Biomarkers 26, 343–353 (2021).

46. Fleury, H. et al. Cumulative defects in DNA repair pathways drive the PARP inhibitor response in high-grade serous epithelial ovarian cancer cell lines. Oncotarget 8, 40152– 40168 (2017).

